# Chest X-ray classification using Deep learning for automated COVID-19 screening

**DOI:** 10.1101/2020.06.21.20136598

**Authors:** Ankita Shelke, Madhura Inamdar, Vruddhi Shah, Amanshu Tiwari, Aafiya Hussain, Talha Chafekar, Ninad Mehendale

## Abstract

In today’s world, we find ourselves struggling to fight one of the worst pandemics in the history of humanity known as COVID-2019 caused by a coronavirus. If we detect the virus at an early stage (before it enters the lower respiratory tract), the patient can be treated quickly. Once the virus reaches the lungs, we observe ground-glass opacity in the chest X-ray due to fibrosis in the lungs. Due to the significant differences between X-ray images of an infected and non-infected person, artificial intelligence techniques can be used to identify the presence and severity of the infection. We propose a classification model that can analyze the chest X-rays and help in the accurate diagnosis of COVID-19. Our methodology classifies the chest X-rays into 4 classes viz. normal, pneumonia, tuberculosis (TB), and COVID-19. Further, the X-rays indicating COVID-19 are classified on severity-basis into mild, medium, and severe. The deep learning model used for the classification of pneumonia, TB, and normal is VGG16 with an accuracy of 95.9 %. For the segregation of normal pneumonia and COVID-19, the DenseNet-161 was used with an accuracy of 98.9 %. ResNet-18 worked best for severity classification achieving accuracy up to 76 %. Our approach allows mass screening of the people using X-rays as a primary validation for COVID-19.

## 2 Introduction

The arrival of novel coronavirus disease (COVID-19), which originated in Wuhan, China, in December 2019 marked the beginning of several unwanted circumstances, such as, economical crisis, loss of life, etc. This disease then spread rapidly from one country to another in a few months. This led to the world health organization (WHO) declaring the COVID-19 outbreak as a global pandemic. Animals are capable of transmitting the coronavirus. But, because of the zoonotic nature of the coronavirus, they have the capability to enter into the human body. Such instances have been witnessed earlier also, i.e. in the case of the severe acute respiratory syndrome coronavirus (SARS-COV) and middle-east respiratory syndrome coronavirus (MERS-COV). They had a major impact on human life leading to many deaths. Some of the clinical symptoms of COVID-19 are muscle pain, shortness of breath, fever, cough, sore throat, headache, fatigue, etc. As soon as a patient experiences signs similar to those of COVID-19, they get tested via standard COVID-19 test. But the results usually arrive within one or two days period. In case of a serious infection, this delay can be harmful. Thus, chest X-rays can prove to be a cheaper, faster, and reliable alternative to standard COVID-19 test. The diagnosis of COVID-19 currently involves various methods that use reverse-transcription polymerase chain reaction (RT-PCR). The sensitivity of RT-PCR is low and there are chances that few results go as a false negative. Hence there was a need for cross-verification examination, such as radiological images.

Clinical images play a vital role in the early detection of the virus. Chest computer tomography (CT) imaging using radiological techniques and X-ray proved to diagnose the initial stages of COVID-19 before the disease spreads further to the lungs causing greater damage. CT is sensitive methods and screening show that the patients have normal CT during the first 2 days after infection. Infection in the lung of the patient is clearly visible after 10 days using CT. Hence X-ray proves faster and better than CT for early screening. Insufficiency of RT-PCR test kits leads to infections being detected late or completely going undetected until an advanced stage is reached.

Machine learning (ML) has wide applications in the medical field, as the machine can replace any professional if it is trained to do a particular task. Hence, machine learning models can be used to assist clinicians to perform tasks with higher speed and accuracy. Deep learning (DL) models (sub-set of ML) are mathematically designed networks that are trained to take certain input and classify it into one of the specified categories. With this advancement in technology, we have deep learning algorithms that are applied to treat several diseases like arrhythmia, skin cancer, breast cancer, pneumonia, and many more. Deep learning models have also been deployed on chest X-ray images and fundus and diagnose diseases. With the advancement in technology, the number of radiologists and hospitals can use deep learning models even when working remotely. In the current COVID-19 pandemic, fast DL models have to be trained and tested to provide timely assistance and accurately produce results. A major constraint so far is the insufficiency of the RT-PCR test and the test has a high cost and it takes relatively more time to produce the results. Deep learning on chest X-ray images can solve such issues and could help in eliminating the disadvantages of RT-PCR and CT scans. Figure 1 is a diagrammatic representation of our fast screening model which classifies the people entering the machine into Infected and Uninfected using a multi-class classification in real-time. Figure 2 shows proposed system flow diagram. A chest X-ray image being passed through a VGG-16 model. The results are bracketed as Normal, Pneumonia, or Tuberculosis. The Pneumonia images are then passed through a DenseNet-161 model and classified into Normal Pneumonia or COVID-19. Then the COVID-19 images are passed through a ResNet18 model and categorized as Severe, Medium, or Mild.

**Fig. 1:**
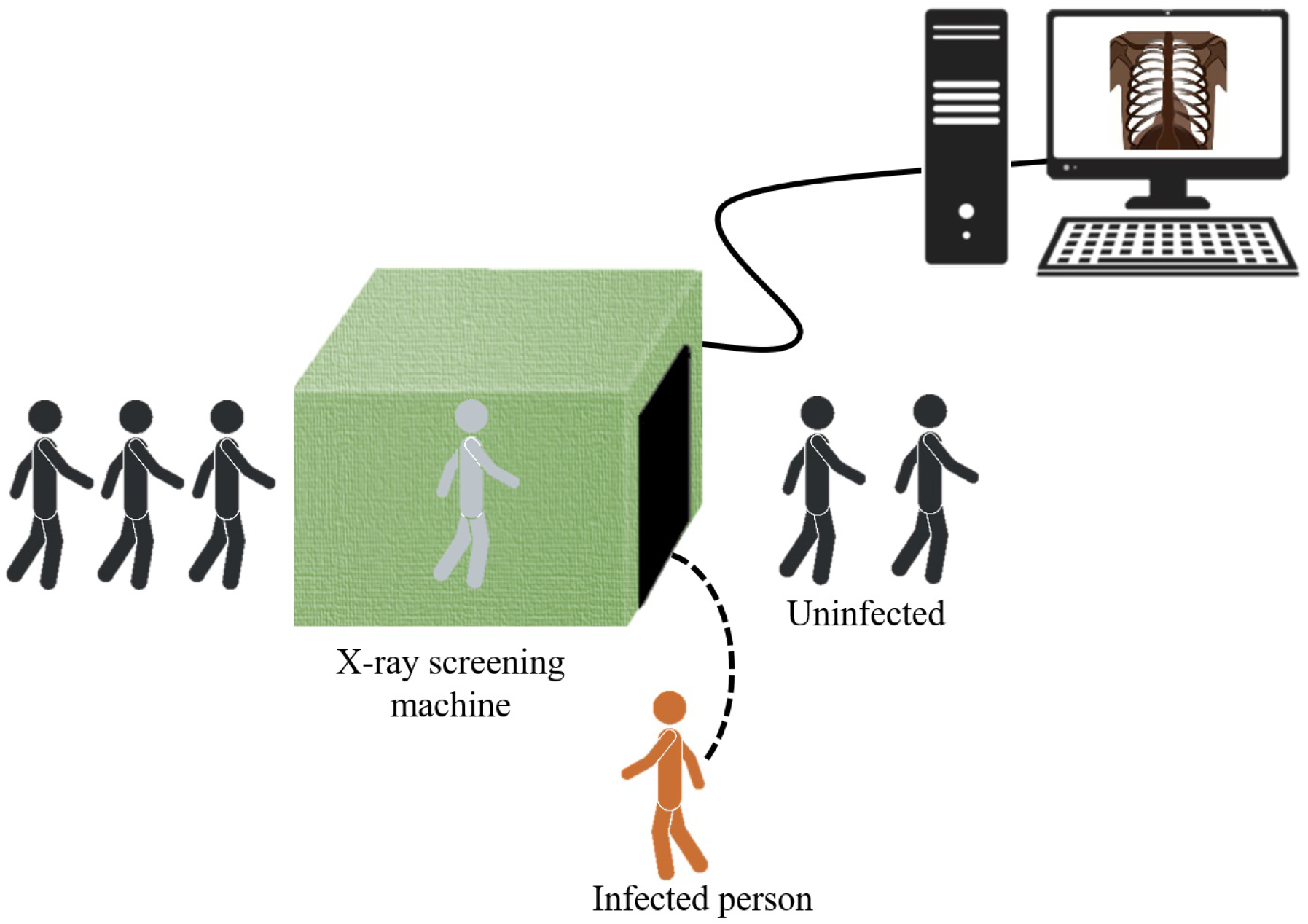
A graphical representation of our fast screening model which classifies the people entering the screening machine into infected and uninfected using a multiclass classification in real-time.

**Fig. 2:**
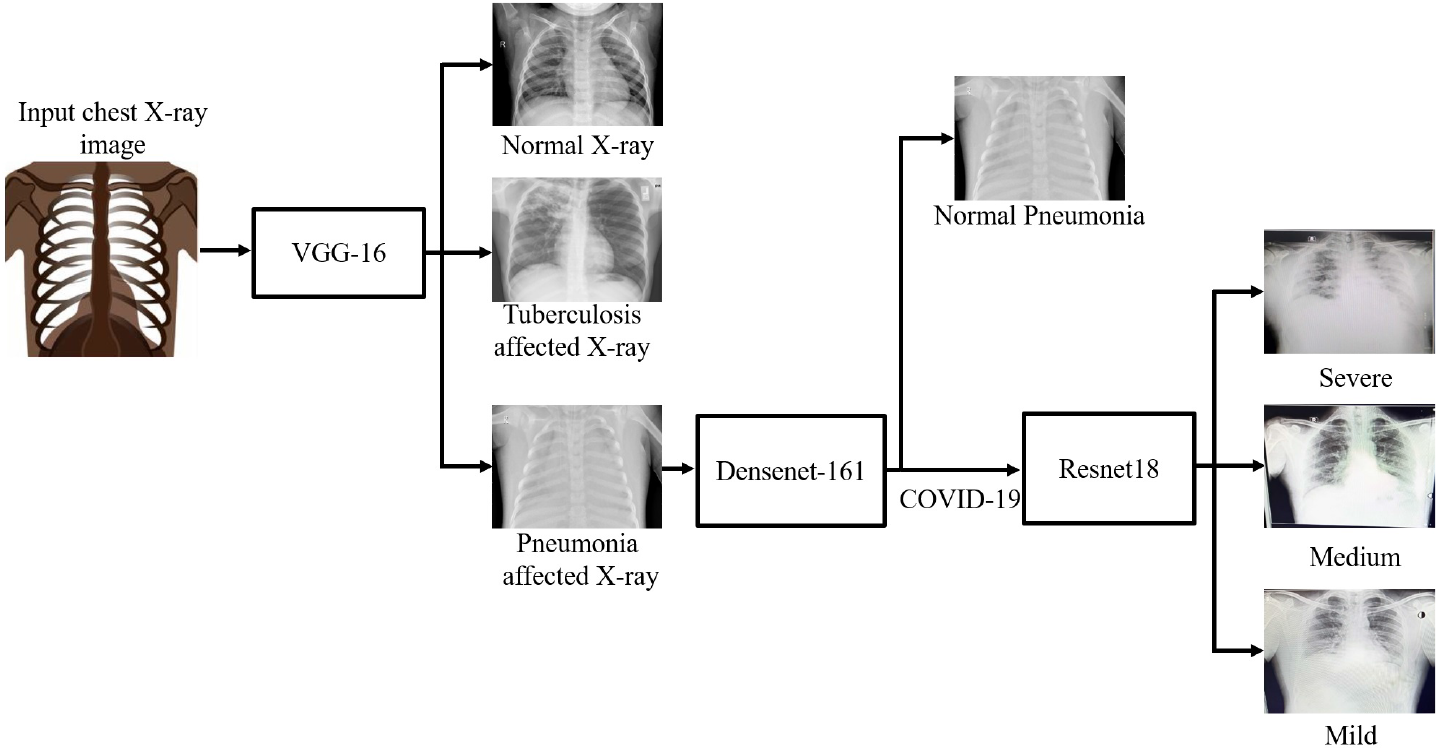
Proposed system flow diagram. The input chest X-ray image is passed through a VGG-16 model and labeled as either normal, tuberculosis, or pneumonia. Further, the pneumonia images are passed through a DenseNet-161 model and categorized as normal pneumonia or COVID-19. The COVID-19 images are passed through a ResNet18 model and classified as severe, medium, or mild.

## 3 Literature review

In recent years, medical imaging has proved to be an effective tool for the diagnosis of a patient [1] with diseases like pneumonia, TB, etc. In the Chest X-ray of a COVID-19 infected patient, ground-glass opacities (GGO)and airspace opacities are observed by Kong *et al*. [2]. Solitary pulmonary nodules (SPN) are discrete, rounded opacity structures that are usually observed during infection in the lungs, on a Chest X-ray. Yoon *et al*. [3] found that the presence of SPNs in a Chest X-ray may indicate the early stage of lung abnormalities. Zhao *et al*. [4] found ground-glass opacities in the patients and vascular dilation in lesions. GGO and consolidation interlobular septal thickening in X-ray images were found by Li *et al*. [5] based on the air bronchogram. Zu *et al*. [6] found that 33 % of the chest CTs had rounded lung opacities. Hemdan *et al*. [7] used deep learning to diagnose COVID-19 using X-ray images using COVIDX-Net. Wang *et al*. [8] used the deep model and the results showed 92.4 % accuracy for datasets of non-COVID, who had pneumonia in the COVID-19 phase. Ioannis *et al*. [9] used Chest X-ray images of 224 COVID-19 positive patients, 700 Pneumonia, and 504 healthy cases. They used VGG-19(a nineteen layer deep convolutional network) and achieved an accuracy of 93.48 %. Narin *et al*. [10] used ResNet50 and deep convolutional neural networks (CNN) for processing the Chest X-ray for diagnosis and achieved an accuracy of 98 %. Sethy *et al*. [11] processed the chest X-ray images with the help of convolutional neural networks (CNN) using ResNet50 and SVM.

## 4 Methodology

A total of 2271 chest X-ray images (895×1024×3 pixels) were obtained from Clinico Diagnostic Lab, Mumbai India. The image was converted from RGB to gray. The resolution of images was reduced to (64×64×1). A total of 1071 images of all three types (i.e. Normal, TB, Pneumonia), split into 70 % training images and the remaining 30 % test images. Input images were fed to Visual Geometry Group (VGG) VGG16. The training dataset consisted of 388 normal images, 500 pneumonia images, and 303 TB images. The pneumonia images consisted of both normal pneumonia and COVID-19 images. The resolution of pneumonia images was kept the same i.e. (64×64×1). The resultant images were sent to the next model which labeled them as normal pneumonia or COVID-19. The dataset used for the training consisted of 1000 images, 500 of which represented pneumonia and 500 represented COVID-19. It used DenseNet-161 and predicted for 233 COVID-19 and 152 pneumonia images. The images that indicated COVID-19 were passed to model for classifying the X-rays based on the severity, which used ResNet-18. The size of the input images was kept at (64×64×1) and a total of 80 images were used for training the ResNet-18. The resultant classes determined on the basis of the severity of COVID-19 were mild, severe, and medium.

As shown in figure 3 We have used a pre-trained network called visual geometry group (VGG) VGG16 for the classification of chest X-rays into Normal, TB, and Pneumonia. VGG-16 is a 19 layer model. The model makes use of Imagenet weights. The input size to the model is (64,64,1) which is fed to a convolutional layer. This convolutional layer increases the channels of the image and gives an output with 64 channels, keeping the dimensions of the image the same as before. The next layer is also a Convolutional layer which gives an output of (64,64,1). After this, we apply max-pooling, which changes the image dimensions to (112,112). The next two layers are convolutional layers, which keep the image channels as (112,112,128). Next, we apply a max-pooling layer, which reduces the image dimensions to (56,56,128). Then, we have a block of 3 convolutional layers and a max-pool layer, whose output is a (28,28,256) image. Next, we have a set of 3 convolutional layers with output dimensions of (28,28,512) and a max-pooling layer with an output of (14,14,512). The final 3 convolutional layers have an output size of (14,14,512) and the max-pooling layer has an output size of (7,7,512). Since we are doing a 3-way classification, we flatten the output of the Max pooling layer and then have a dense layer with 3 neurons. The learning rate was kept at 0.001 with Adam as the optimizer. We trained the model for 12 epochs.

**Fig. 3:**
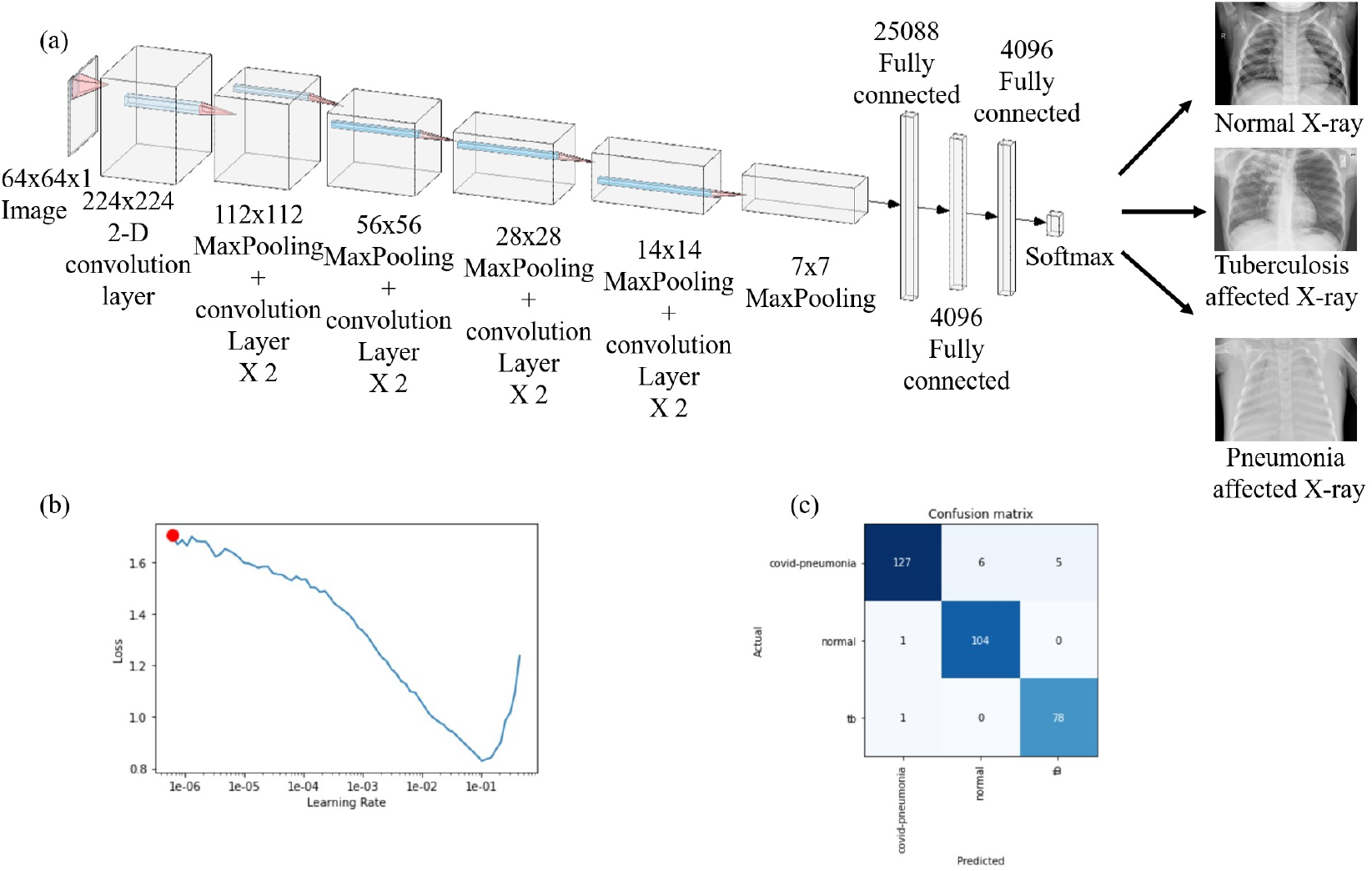
Configuration and performance of VGG16 (a) Model of VGG16. The input size of the image is 64×64×1. After this, the image passes through a 2D convolution layer with dimensions 224×224. Then it passes through the next layer with dimensions 112×112. After this, it passes to consecutive convolution layers along with MaxPooling with dimension changes 56×56, 28×28, and 14×14. Then it goes to a Max Pooling layer with dimension change 7×7. After a series of 2500, 4096, and 4096 fully connected layers, the X-rays are classified into Normal X-ray, Tuberculosis affected X-ray, and Pneumonia affected X-Ray. (b) The graph of the learning rate versus a loss in the range of 0.8 and 1.7. (Red dot shows selected learning rate) (c) A total of 322 images were tested. 127 were correctly labeled as COVID-Pneumonia, 104 were correctly labeled as Normal and 78 were correctly labeled as Tuberculosis.

For the next step in binary classification, we have used the DenseNet-161 pre-trained model (fig. 4). An image of (64,64,1) is fed to the model. We have a total of 4 Dense blocks with 3 transitional layers in between. Each transitional layer consists of a 1×1 convolutional layer and an average pool layer of size (2,2) with a stride of 2. Each dense block consists of 2 convolutional layers. The first block has a 1×1×6 layer and a 3×3×6 layer. The second block has a 1×1×12 and a 3×3×12 layer. The third block consists of a 1×1×48 and a 3×3×48 layer. The final dense block consists of a 1×1×31 and a 3×3×31. The final classification layer is a 7×7 global average pool layer and finally a softmax layer of 2 neurons. The learning rate was chosen to be 0.002 and the model was trained for 15 epochs. We have performed a 3-way classification between severe, mild, and medium severity of the COVID-19 X-ray images. As shown in figure 5, pre-trained ResNet-18 was used for this purpose. The input image for this model is a 64×64×3 image. This model consists of 5 convolutional blocks. The first convolutional block consists of a 7×7×64 kernel. In the next convolutional block, we have a 3×3 max pool layer with a stride of 2. For the second block, we have two (3×3×64) blocks. The third, fourth, and fifth convolutional blocks have the filter dimensions of 128,256,512 respectively with each of the kernel sizes as 3×3. There is an average pooling layer after these and a fully connected layer with 3 neurons for a 3-way classification. The learning rate used was 0.002 and the model was trained for 15 epochs.

**Fig. 4:**
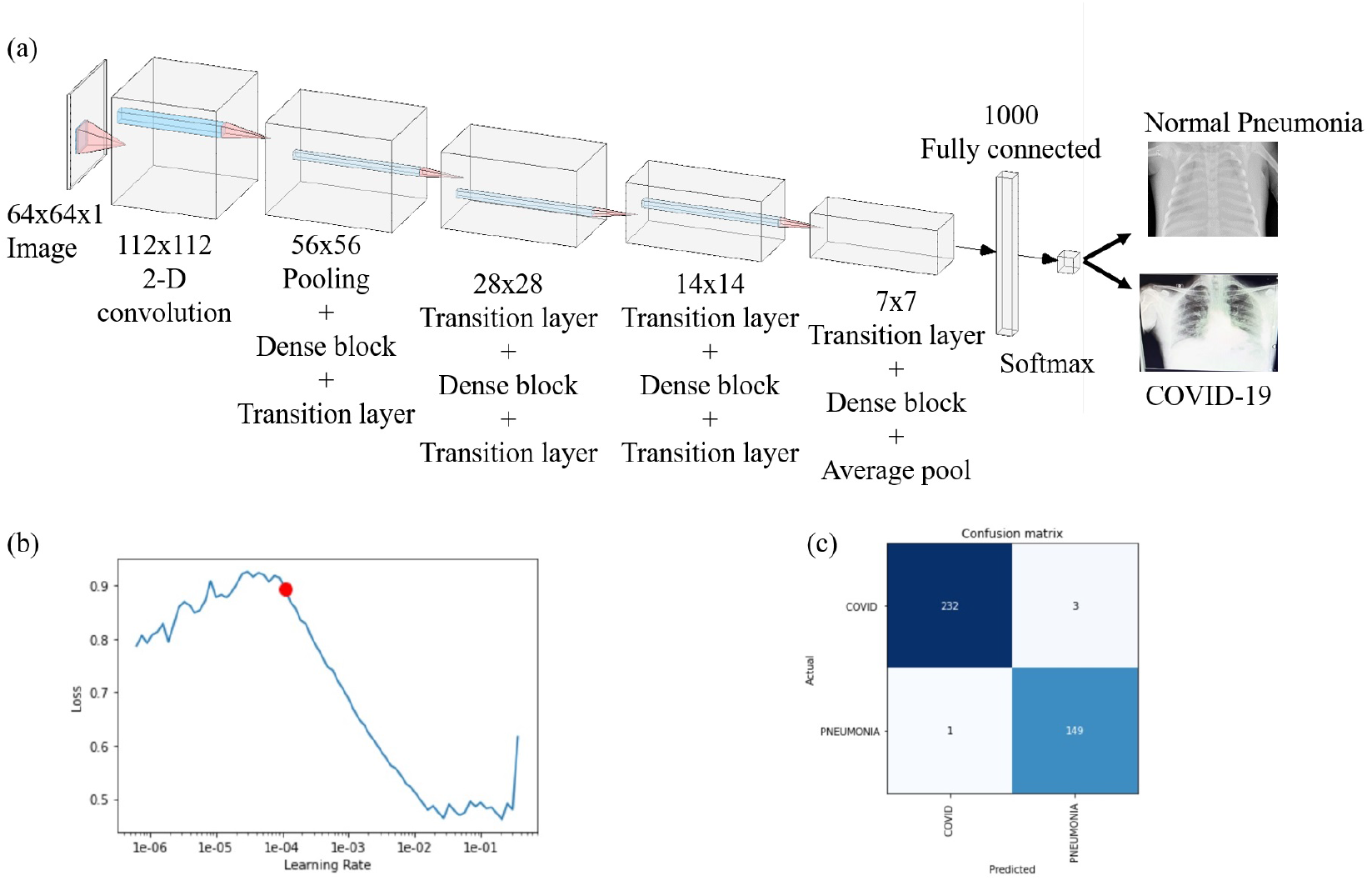
Configuration and performance of DenseNet-161 (a) Model of DenseNet-16. The input size of the image is 64×64×1 after which the image passes through a 2-D convolutional layer with dimensions 112×112. Then it passes through the next layer with dimensions 56×56 After passing through the consecutive layers the dimensions change from 56×56 to 28×28, 14×14, and finally 7×7. After 1000 fully connected layers, the X-rays are classified into normal pneumonia and COVID-19. (b) The graph of the learning rate versus a loss in the range of 0.5 to 0.9. (Red dot shows selected learning rate) (c) Confusion matrix of the output for DenseNet-161. A total of 235 images were tested, out of which 232were correctly classified as COVID, and 3 were found to be wrongly classified. From 150 images, 149 were correctly categorized as pneumonia whereas 1 was wrongly classified.

**Fig. 5:**
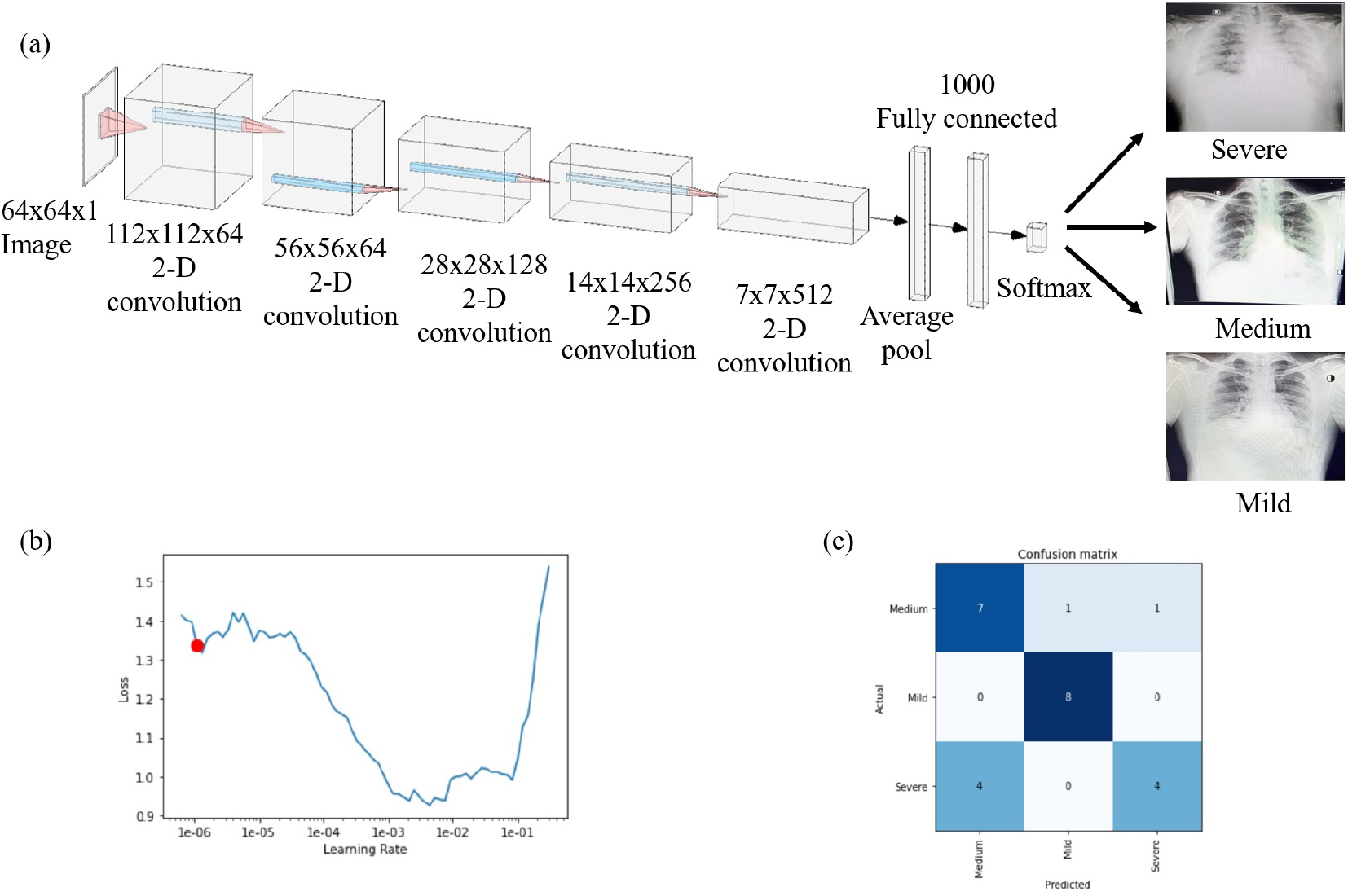
Configuration and performance of ResNet-18 (a) Model of ResNet-18. The input size of the image is 64×64×1. After this, the image passes through a 2D convolution layer with dimensions 112×112×64. Then it passes through the next layer with dimensions 56×56×64. After this, it passes to consecutive 2D convolution layers with dimension changes 28×28×128, 14×14×256, and 7×7×512. After 1000 fully connected layers, the X-rays are classified into Severe, Medium, and Mild. (b) The graph of the learning rate versus a loss in the range of 0.9 and 1.4. (Red dot shows selected learning rate) (c) Confusion matrix of the output for ResNet-18. A total of 25 images were tested, out of which 7 were correctly labeled as Medium, 8 were correctly labeled as Mild and 4 were correctly labeled as Severe.

## 5 Results and discussion

For table 1, NPT stands for Normal, Pneumonia, and Tuberculosis. RC stands for Regular Pneumonia and COVID Pneumonia. SMm stands for Severe, Medium and mild. To validate the results, we first trained the VGG16 network using 970 labeled images via supervised learning. 270 images were used to test the pre-trained VGG16 network which resulted in an accuracy of 98 % between pneumonia infected, TB infected, and non-infected X-rays.

**Tab. 1:** Comparison of % accuracy Results with respect to different Deep learning networks

|  | NPT | RC | SMm |
| --- | --- | --- | --- |
| Alexnet | 50 | 70 | 50.4 |
| Resnet-18 | 72.3 | 78.2 | 76 |
| Resnet-50 | 60.4 | 89.5 | 60 |
| VGG-13 | 70.2 | 87.8 | 40.9 |
| VGG-16 | 95.9 | 78.9 | 40 |
| DenseNet-161 | 89.8 | 98.9 | 54.2 |
| Densenet-169 | 78.6 | 94.7 | 65.8 |
| Densenet-201 | 85.9 | 93.2 | 60 |

A total of 1000 images were chosen out of which 500 were regular pneumonia X-ray images and the remaining 500 were COVID infected X-ray images. These images were used to train the DenseNet-161 Network and the ratio of training and testing was 70: 30 and we found that with 99 % accuracy, we can segregate pneumonia from COVID infected chest X-rays. It has been quite difficult for expert radiologists to differentiate pneumonia from COVID-infected X-rays and making it possible through deep learning was a very big achievement. On the request of radiologists, we also tried to segregate COVID X-rays based on the severity of the infection into 3 subclasses viz. mild, medium, and severe. For this, we only had 80 labeled images of each subclass. These images were used to train and test on multiple networks and we found that out of all the ones that were trained and tested, ResNet18 performed well with an accuracy of up to 76 % to classify between the three subclasses. Overall, there is still a scope of improvement in accuracy for classifying the COVID-19 infected X-rays into three different classes.

Figure 3 (b) depicts the graph of the learning rate versus a loss in the range of 0.8 and 1.7. The red dot shows the selected learning rate. Figure 3 (c) shows that a total of 322 images were tested. Out of these images,127 were correctly labeled as COVID-Pneumonia, 104 were correctly labeled as Normal and 78 were labeled as Tuberculosis. The graph of the learning rate versus a loss in the range of 0.5 to 0.9 can be seen in figure 4 (b). The red dot shows the selected learning rate. Figure 4 (c) demonstrates the Confusion matrix of the output for DenseNet-161. A total of 235 images were tested, out of which 232 were correctly classified as COVID, and 3 were found to be wrongly classified. From 150 images, 149 were correctly categorized as pneumonia whereas 1 was wrongly classified. Figure 5 (b) illustrates the graph of the learning rate versus a loss in the range of 0.9 and 1.4. The red dot shows the selected learning rate. The confusion matrix of the output for ResNet-18 is observed in figure 5 (c). A total of 25 images were tested, out of which 7 were correctly labeled as Medium, 8 were correctly labeled as Mild and 4 were correctly labeled as Severe.

For classification, of X-ray into 3 types i.e. TB, Pneumonia, and Normal we used AlexNnet first. The disadvantage of AlexNet was that it had a lesser number of layers and thus the accuracy obtained was 50 %. Secondly, we trained and tested ResNet-50 model which comparatively showed a better accuracy of 60.4 % but the time it took to train was too long and the model was too large, so we tried VGG-13 and obtained an accuracy of 70.2 % and then VGG-16 was also trained and tested which gave by far the best results. VGG16 gave an accuracy of by far the highest which is 95.9 %. Other models like ResNet-18, DenseNet-161, DenseNet-169, DenseNet-201 were trained and tested but did not show any improvement in the accuracy. For further classification of images into Pneumonia or COVID-19, we started first with AlexNet since the model is smaller in size. We got 70 %. We then trained a ResNet-50 model which gave us an accuracy of 89.5%. We looked for much bigger models to train and hence used DenseNet-201 which gave us an accuracy of 93.2 % but the model size was quite large and the training took long. Other models like ResNet-18, DenseNet-161, DenseNet-169 were trained and tested but did not show any improvement in the accuracy. Hence, we tried a smaller version of the DenseNet, i.e. DenseNet-161 which gave us an accuracy of 98.9 %. For further classification, of COVID-19 positive X-ray into 3 types i.e. mild, severe, and medium we used AlexNet first. The disadvantage of AlexNet was that it had a lesser number of layers and thus the accuracy obtained was 50.4 %. Secondly, we trained and tested the DenseNet-161 model which gave an accuracy of 54.2%. Further, we trained DenseNet-169 which comparatively showed a better accuracy of 65.8 %. ResNet-18 gave an accuracy of by far the highest of 76 %. Other models like ResNet-50, DenseNet-161, DenseNet-201 were trained and tested but did not show any improvement in the accuracy.

### 5.1 Comparison with existing literature

Table 2 shows that, some of the reported research groups have used X-ray reports whereas some have used CT reports to attain COVID-19 results. We have sequentially used 3 methods to attain the required results. The accuracy of the VGG-16 model, DenseNet-161 model, and ResNet 18 gave us an accuracy of 95.9 %,98.9 %and 76 % respectively. We achieved the highest accuracy which can further be improved.

**Tab. 2:** Comparison with other methods. Most of the methods classify X-rays only into pneumonia, COVID, and normal, our method further classifies COVID-19 into 3 categories.

| Method | Type of images | Number of cases | Method Used | Accuracy (%) |
| --- | --- | --- | --- | --- |
| Ioannis *et al.* [9] | Chest X-ray | 1428 | VGG-19 | 93.48 |
| Wang *et al.* [8] | Chest X-ray | 13645 | COVID-Net | 92.4 |
| Sethy *et al.* [11] | Chest X-ray | 50 | ResNet50+SVM | 95.38 |
| Hemdan *et al.* [7] | Chest X-ray | 50 | COVIDX-Net | 90 |
| Narin *et al.* [10] | Chest X-ray | 100 | Deep CNN | 98 |
| Ying *et al.* [12] | Chest CT | 1485 | DRE-Net | 86 |
| Zheng *et al.* [4] | Chest CT | 542 | Unet+3D<br>Deep Network | 90.8 |
| Xu *et al.* [6] | Chest CT | 618 | ResNet+<br>Location Attention | 86.7 |
| Proposed<br>Method | Chest X-ray | 22 | VGG-16 | 95.9 |
|  |  |  | DenseNet-161 | 98.9 |
|  |  |  | ResNet-18 | 76 |

### 5.2 Discussions

In the future, the chest X-ray machines can itself act as a source of detecting COVID-19 and further diagnosis can be carried out using RT-PCR, saving the hard-earned money of patients. Furthermore, X-ray machines are well available in bulk, and in most of the regular hospitals, the X-ray machine is an integral part of the setup and it will be easy to make this available in local hospitals as well. Adding to that, X-ray machines require less maintenance compared to RT-PCR in terms of their reagents, and hence the cost of operation is comparatively low. There are some side effects of X-ray scanning, such as people carrying metals and pregnant women, for such cases, we recommend that X-rays should not be used, otherwise mass-screening through X-ray is possible.

## 6 Conclusion

Deep learning can be a very useful tool in the medical industry for the detection of diseases just by processing the images of the chest X-rays and feeding it as an input to the model. The chest X-rays are initially classified into different classes such as normal, pneumonia, tuberculosis, and COVID-19. After distinguishing normal pneumonia from the COVID positive patient X-ray, as a proof of concept, we have shown that COVID-19 X-rays can be classified based on the severity of the COVID-19 into different sub-units such as severe, mild, and medium stage.

Mass screening of people for detection of COVID-19 can be done effectively with our proposed model. It will help yield faster and accurate results and will be cost-effective as compared to the conventional RT-PCR method. This approach can be implemented at the local level and in the rural areas where adequate facilities are absent.

## Data Availability

Data can be made available in the public domain on request

## acknowledgements

Authors would like to thank Dr. Palande from Clinico Diagnostic Lab for providing the images and Dr. Madhura Mehendale for their support in labeling the supervisory dataset.

## Compliance with Ethical Standards

### Involvement of human participant and animals

This article does not contain any studies with animals or Humans performed by any of the authors. All the necessary permissions were obtained from the Institute Ethical Committee and concerned authorities.

### Information about informed consent

Informed consent was not required as there were no participant

